# Shining light on the dark matter of pertussis: evidence for an asymptomatic carriage state from a longitudinal cohort of mother/infant dyads

**DOI:** 10.1101/2025.01.20.25320623

**Authors:** Christian E Gunning, Christopher J Gill, Pejman Rohani

## Abstract

Pertussis remains an enigma partly due to the uncertain impact of asymptomatic cases on transmission. Contributing to this knowledge gap is a lack of high-quality disease surveillance, particularly in those low- and middle-income countries that experience high disease burdens. Here we present analyses based on our prospective longitudinal surveillance of a rolling cohort of 1,315 mothers and their newborn infants in Lusaka, Zambia across 2015 (8,704 unique study visits). We detail the timing, duration, and intensity of qPCR-based IS481 signals in individual subjects and within mother/infant dyads. We find that IS481 signal strength: A) in mothers predicts contemporaneous and future IS481 detections in infants, B) in infants predicts, to a lesser extent, detections in mothers, and C) predicts respiratory symptoms in infants but not mothers. We profile a subgroup of 50 infants and 54 mothers who displayed evidence of persistent colonization (median duration 8 weeks) wherein most mothers were entirely asymptomatic. We also include a critical assessment of qPCR test reliability across IS481 signal strengths. Our results demonstrate the routine occurrence of long-duration, mild and minimally symptomatic pertussis infections and suggest that pertussis transmission occurs between minimally symptomatic mothers and their newborn infants.

## Introduction

The resurgence of *Bordetella pertussis* in a number of countries with high routine immunization coverage (1) has called into question key assumptions about the transmission, immunology, and evolution of this highly contagious respiratory pathogen (2, 3, 4, 5, 1). Chief among these questions is the epidemiological importance of mild and asymptomatic pertussis infections (6, 7, 8, 9, 10). Indeed, despite widespread recognition that mild pertussis is common (2), especially amongst adults (11, 12), its contribution to overall disease transmission and persistence remains poorly characterized (13, 6, 14).

Mild and asymptomatic pertussis has sometimes been considered the “dark matter” of pertussis transmission, owing to its elusive yet potentially significant impacts (9, 15). Prior to modern molecular surveillance, reliable detection of mild pertussis infections was challenging (16, 17, 18) and, in the pre-vaccine era, the epidemiological role of mild pertussis was largely ignored (19, 20, 21). As vaccination rates rose worldwide across the 20th Century, pertussis morbidity and mortality dropped sharply (22, 23, 24, 4). Yet local pertussis elimination has rarely been achieved, even in highly-vaccinated populations, unlike many similar childhood diseases such as measles, mumps, and rubella. Furthermore, the relative protection afforded by vaccination against transmissible infection versus carriage remains a subject of active debate, as does the precise epidemiological role of mild and asymptomatic pertussis infections (1). Consequently, understanding the determinants of pertussis transmission and persistence remains a pressing global health concern and an area of active public health research (25).

In recent decades, novel molecular surveillance tools have yielded remarkable insights into global pertussis dynamics, from phylodynamics (3, 26) to pathogen evolution (27, 28, 29, 30). Notably, qPCR-based testing for pertussis can detect mild pertussis much more reliably than previous culture-based methods (31, 32, 33). The most common of these tests targets a repetitive genetic sequence, IS481, that occurs at copy numbers in excess of 200 in *B. pertussis*. While IS481’s high copy number allows for high qPCR test sensitivity (<1 bacterium per reaction well), the interpretation of weak qPCR signals (i.e., low target counts) has been the subject of considerable debate. Some have argued for using additional tests (i.e., culture or serology) to ‘confirm’ qPCR results (34). Yet the discriminatory value of additional testing is far from clear at present given the low sensitivity of culture and the differing time scales of serology-based tests (35, 36, 34, 37). Furthermore, available tests struggle to reliably distinguish between *B. pertussis* presence, persistent colonization, and transmissible infection (38, 39). In this context, we view qPCR results as highly informative on their own, especially in the context of population and longitudinal surveillance.

Deployed at scale, qPCR offers the potential for a deeper understanding of when and where mild pertussis is most likely to occur, and how these infections contribute to local and regional disease dynamics. At present, however, high-income countries are responsible for the bulk of routine molecular pertussis surveillance (15). As these countries typically also experience the lowest burdens of most infectious diseases, such concentrated surveillance means that the best-studied populations may offer hard-won yet modest epidemiological insights.

Our 2015 Southern Africa Mother/Infant Pertussis study (SAMIPS) sought to bridge this gap through the use of targeted, high-resolution pertussis surveillance in a low-resource urban community (40, 41). Conducted in Lusaka, Zambia across one calendar year, SAMIPS followed a cohort of mothers and their young infants across these infants’ first 3-4 months of life. Each mother/infant dyad attended regularly-scheduled visits at a local public health clinic (PHC), where self-reported respiratory symptoms were recorded and nasopharyngeal (NP) samples were collected. At these visits, routine vaccinations and well-child care were also administered and recorded. Overall, our partnership with the local PHC and establishment of an on-site molecular laboratory enabled us to (I) conduct prospective longitudinal surveillance of the SAMIPS cohort and (II) link detailed clinical records with qPCR testing of nasopharyngeal (NP) samples. Critically, we designed SAMIPS to minimize selection biases by sampling on a fixed schedule rather than responding to apparent symptoms. This framework allowed us to assess the burden of symptomatic pertussis, as is generally done when only testing children who present with a clinical syndrome typical of pertussis, as well as asymptomatic infections. By sampling mothers and infants on the same schedule, we also gained insights into the epidemiology and clinical presentation of pertussis within mother/infant dyads.

Our previous work in this cohort has profiled a small number of the most sick infants (40), inspected the timeliness of DTP vaccine uptake in cohort infants (41), and described antibiotics administration across the cohort (42). We also quantified evidence for pertussis infection (EFI) across the full cohort, where we observed a strong correspondence of EFI between mothers and their infants (10). To our surprise, we also found a strong correspondence between EFI and mild symptoms (cough and/or coryza alone) in infants as well as mothers, suggesting the common occurrence of mild pertussis in infants. We were also intrigued by the detection of apparently asymptomatic pertussis among a subset of infants with strong EFI (10).

Our previous findings underscored the epidemiological importance of mild and asymptomatic pertussis, especially in low-resource settings, and highlighted the prospect of weaker IS481 qPCR signals as sensitive indicators of pertussis infections in both populations and individuals. Notably, we identified numerous individuals with strong evidence of pertussis infection within a single year in one community in Lusaka, despite the complete absence of any official pertussis case reports in all of Zambia since 2009 (43). In short, our results suggested a systematic inability to detect ongoing pertussis transmission at a regional and national level.

Our previous work, however, left several key questions unaddressed. Here we explore the detailed time-course of qPCR tests and respiratory symptoms within individual subjects and within mother/infant dyads. We also critically examine the fidelity of our qPCR-based pertussis testing in order to assess the reliability and robustness of our results. Finally, we profile a group of subjects with evidence of persistent colonization. Altogether, our results offer granular insights into otherwise invisible pertussis dynamics in this at-risk population of mothers and their infants.

## Results

### Study Overview

SAMIPS was designed to enroll, across one calendar year, all newborns and their mothers in Chawama compound, a low-resource peri-urban community with a population of approximately 150,000 people at the time of the study (2015). The study was conducted at the single no-cost public health clinic available to Chawama residents, where subjects were observed at enrollment and (ideally) at six additional scheduled study visits across infants’ first 100 days of life (plus *ad hoc* sick visits). At each visit, we recorded vital signs, antibiotics prescriptions, the administration of routine vaccinations (including DTP doses 1-3), and self-reported and observed respiratory symptoms. Critically, we also collected nasopharyngeal (NP) samples at each visit that were retrospectively tested for pertussis (and other common respiratory pathogens) via qPCR at our lab located at Lusaka’s University Teaching Hospital (UTH).

We lack maternal vaccination records, but note that WHO estimates indicate DTP3 coverage in excess of 80% back to 1985 (44). This fact, along with the potential of widespread under-reporting of transmission in pertussis cases in LMICs, leads us to believe that few (if any) mothers were immunologically naive at the beginning of the study. We also note that genetic sequencing of pertussis was not conducted due to its reliance (at the time) on culturing. For additional details, including cohort demographics, see Gill et al. (40), Gunning et al. (41) and Gill et al. (10).

Ultimately, SAMIPS enrolled nearly 2,000 mother/infant dyads and conducted qPCR tests on over 19,000 NP samples. We focus here on the subset of 1,315 dyads who attended at least 4 study visits, for a total of 17,408 unique NP samples across 8,704 dyad visits (including 410 unscheduled visits).

### qPCR: detection versus signal strength

For clinical pertussis diagnosis, IS481 *Ct* < 35 is a frequently used threshold (15), yet experimental variation between individual tests, plates, and labs precludes a direct estimate of sample quantity from *Ct* value in the absence of rigorous controls (45, 46, 47). Accordingly, we use the detection and relative quantification of IS481 here to focus on disease surveillance and epidemiological inference. In this context, we first classify IS481 qPCR tests as either detections (i.e., *Ct* < 45) or non-detections (N.D.). As before (10), we further categorize detections by signal strength into three arbitrary groups: weaker (*Ct ≥* 43), intermediate (43 > *Ct ≥* 40), and stronger (40 > *Ct*). *A priori*, we expect that stronger signals are more reliably detected and are more predictive of symptomatic illness, carriage, and disease transmission. We proceed with the assumption that all IS481 detections are *potentially* informative, and directly interrogate the information content of qPCR tests by their signal strength.

Overall, we observe detections in 9% of NP samples (N=1,540, Table 1bA). Most of these detections consist of weaker IS481 signals (median *Ct* = 43.2); *Ct* < 40 in 10.2% of detections (N=159), and *Ct* < 35 for 1.7% (N=26). In Figure 1A, we show the frequency of IS481 detections by *Ct* value for mothers vs infants. Notably, stronger IS481 signals (*Ct* < 40) are observed much more frequently in infants (10% of detections) vs mothers (3.7%). We also summarize the strongest observed IS481 signal by subject and dyad (Figure 1B), again highlighting a long tail of stronger IS481 signals in infants. In Figure 1C, we show the number of mothers, infants, and dyads with one or more detections across their course of study participation. In addition, Table 1B tabulates the frequency of co-detection within a dyad’s visit (OR=4.9, 95%CI: 4.1-5.9). In all cases, the observed overlap between infants and their mothers is much higher than expected by random chance, illustrating how detections tend to cluster within dyads.

**Table 1:**
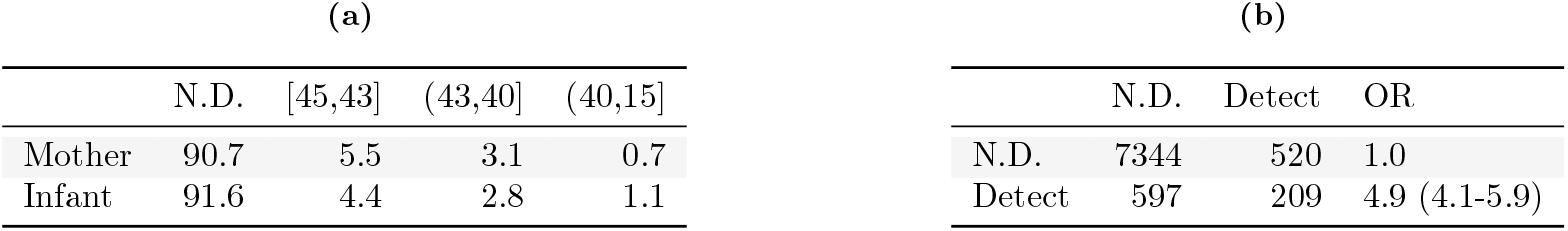
a) Percent of samples by IS481 signal strength across 8,704 dyad visits. b) Odds ratio of co-detection (OR +95% CI; rows show mothers, columns show infants, N=8,704 unique dyad visits).

**Figure 1:**
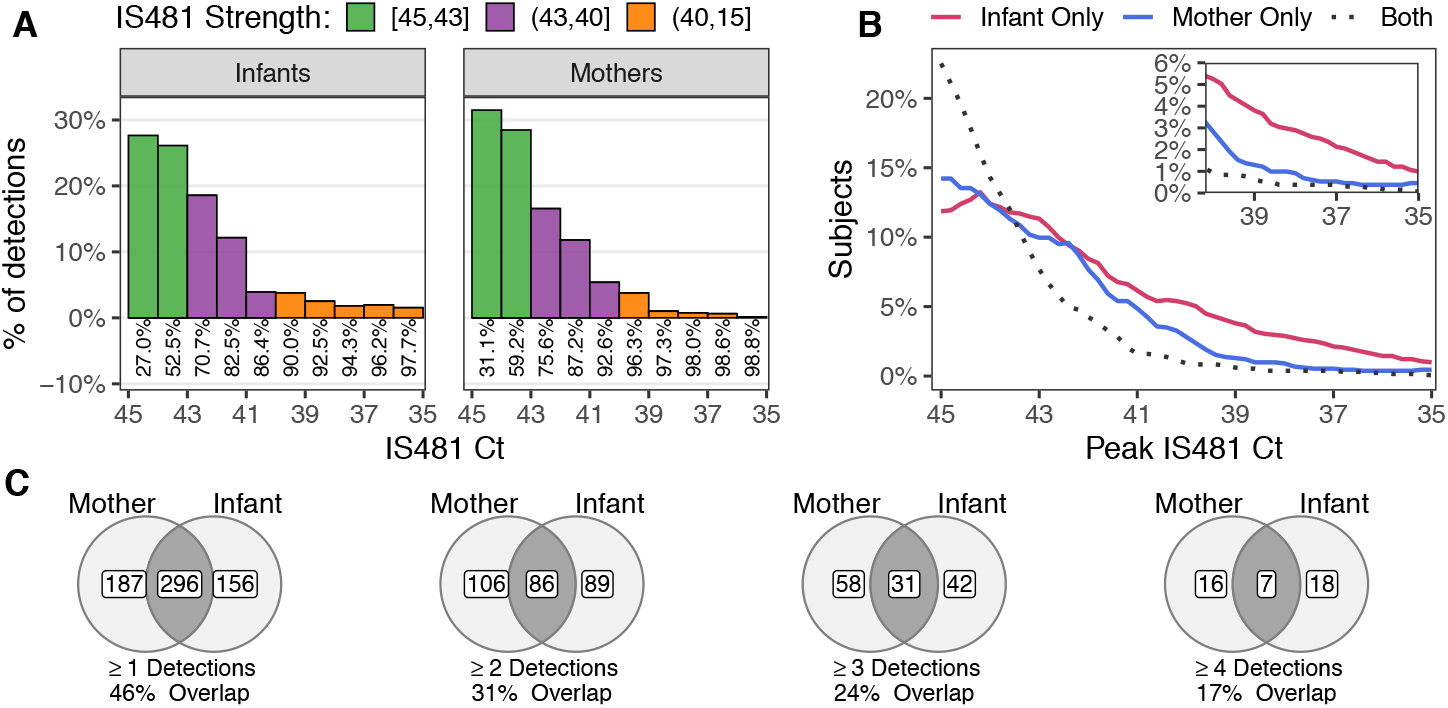
Overview of qPCR test results showing A) frequency of IS481 *Ct* values (by visit), B) frequency of peak IS481 values (by subject and dyad), and C) number of IS481 detections across full study (by dyad). A) Intermediate and stronger IS481 signals (color) were much more common in infants than mothers (inset numbers show cumulative frequency from left to right; *Ct* < 35 not shown). B) More infants exhibit stronger peak IS481 values, while dyad members together frequently exhibit weaker peak signals (dashed line). C) Venn diagrams show the frequency of subjects with *≥* 1 (left) to 4 (right) detections (*Ct* < 45), stratified by infants, mothers, and both dyad members (shaded region).

### Study Timeline

We employed a rolling enrollment of the study cohort such that each dyad’s participation spanned a portion of the full study period. Figure 2A illustrates the study participation of six example dyads, highlighting infrequent unscheduled visits along with the sometimes uneven timeline of scheduled visits relative to the intended schedule. This figure also highlights the potential clumping of detections over subjects’ study participation, a point that we return to later. Figure 2B shows the number of NP samples (and associated qPCR tests) by calendar week and IS481 signal strength, highlighting the changing cohort size across the study period. Figure 2C shows the frequency of NP samples by IS481 signal strength from a generalized additive model (GAM) that accounts for the variable cohort size. As previously described (10), we observe a burst of stronger IS481 signals in June 2015 that is preceded by a more gradual surge of weaker signals in the previous six weeks.

**Figure 2:**
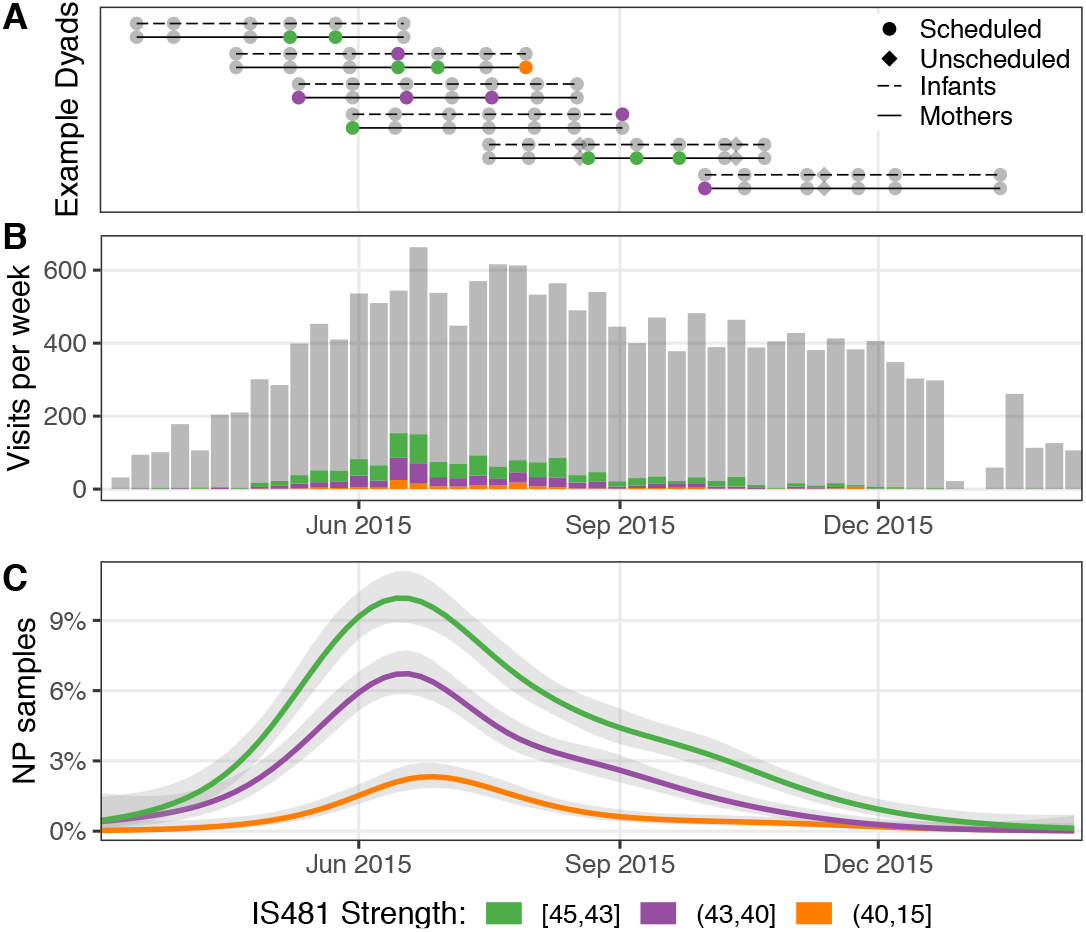
Study timeline. A) Timelines of select dyads (mother/infant pairs) showing IS481 qPCR signal strength. Dyad study visits were scheduled at approximately two week intervals (median 16 days). B) Subject visits per week, stratified by IS481 signal strength (color, grey shows N.D.). C) Estimated frequency of IS481 detections by signal strength over time (GAM, mean +95% CI).

### Experimental re-testing

During the course of qPCR testing, an opportunistic subset of 1,468 NP samples was tested a second time, including 399 initial detections and 1,069 initial N.D.s. In this subset we observed a very low probability of follow-up detection given initial N.D. of 2.3%, suggesting a low false positive rate. By contrast, in samples where initial qPCR results (*Ct*_0_) were stronger (*Ct*_0_ < 40), we found a high probability of confirmation of 86%. In total, this subset revealed a relative risk of confirmation of 21.9 (95% CI: 14.7-32.6, see Table 2), indicating very high overall repeatability of IS481 detection.

**Table 2:** Relative risk of confirmation (columns) given initial detection (rows) in an opportunistic subset of 1,468 NP samples that were retested.

|  | N.D. | Detect | RR |
| --- | --- | --- | --- |
| N.D. | 1044 | 25 | 1.0 |
| Detect | 195 | 204 | 21.9 (14.7-32.6) |

These results prompted us to conduct a systematic evaluation of IS481 qPCR reliability as a function of signal strength. To do this, we re-tested a random subset of NP samples. We stratified by initial qPCR strength and across mothers and infants, drawing 10 NP samples per strata across 6 groups of *Ct*_0_ (129 total retested NP samples). As expected, we found no differences between mothers and infants. In Figure 3A, we show the re-test confirmation frequency by *Ct*_0_ strata: here, the proportion of confirmed detections rose rapidly from approximately 25% for *Ct*_0_ > 42 to 58% for 41 > *Ct*_0_ > 40 and 89% for 40 > *Ct*_0_. We also illustrate the overall detection frequency for the full library of qPCR results (8.8%, dashed line), which we henceforth employ as a baseline “null frequency” of detection, representing a conservative upper bound on the qPCR false-positive rate.

**Figure 3:**
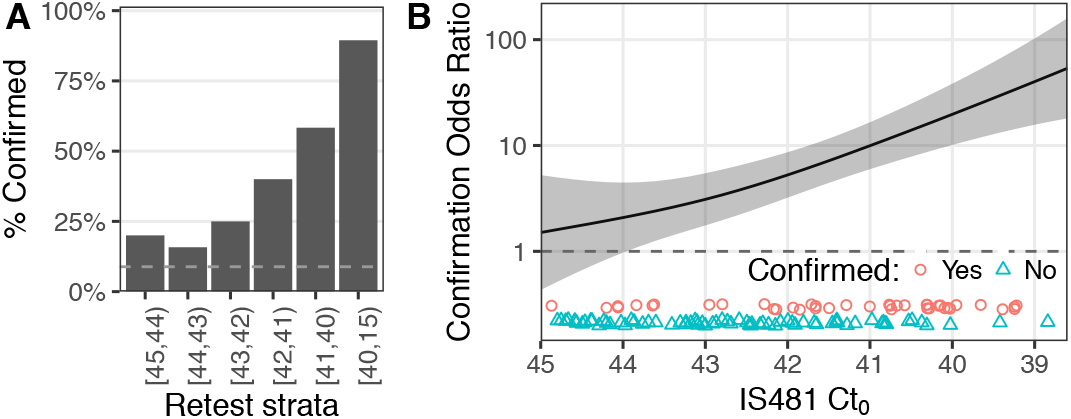
In a stratified random subsample of 122 NP samples with initial detections, re-test confirmation frequency increased with initial IS481 signal strength. A) shows confirmation frequency by subsample strata. B) Odds ratio of confirmation by IS481 signal strength (OR relative to null frequency of detection across full sample library; mean and 95% CI estimated from a binomial GLM). Points show initial IS481 *Ct* stratified by confirmation (shape, color).

In Figure 3B, we consider *Ct*_0_ as a continuous predictor of qPCR re-test confirmation. We use a GAM to predict the odds of confirmation, and then normalize the resulting prediction by the null odds of detection (above) to obtain an estimated mean odds ratio of confirmation (ORoC). Notably, the ORoC’s 95% CI ceases to overlap with 1 just below *Ct* = 44, showing that the odds of confirmation exceed the null odds. At *Ct*_0_ = 43, the estimated ORoC increases to 3.1 (95% CI: 1.8-5.5) and at *Ct*_0_ = 40 to 19.7 (95% CI: 10.1-38.3). In summary, confirmation was more likely than expected by chance alone (i.e., than the null frequency of detection observed in the full NP sample library) across the full spectrum of *Ct* values, including even weaker signals.

We note that the above results are consistent with the expected stochastic variation intrinsic to qPCR. For example, assuming a realistic PCR efficiency of 1.7, we could expect a *Ct* of 42 given 10 targets per sample (i.e., at the theoretical limit of quantification; see Fig. 5 in Ruiz-Villalba et al. (47)). We would also expect Poisson sampling variation in per-reaction target number to yield an approximately continuous decrease in re-test confirmation probability with increasing *Ct*, as observed here (e.g., see Fig. 9 in Ruiz-Villalba et al. (47)).

### Quantifying the information content of IS481 signal strength

The preceding results bolster our confidence that all qPCR tests are *potentially* informative, particularly for epidemiological and public health surveillance where sensitivity often takes precedence over specificity. With this in mind, we next assess the information content of qPCR tests within the longitudinal context of both individuals and mother/infant dyads.

### Time to first detection: reciprocal impacts within dyads

We first employ a survival analysis framework to investigate how the time to initial detection in one dyad member (the *ego*) is affected by preceding IS481 signals within their *alter* (i.e., the ego and alter represent *either* mother and infant, or infant and mother). Here, model covariates include the ego’s identity (infant versus mother) and the alter’s IS481 signal strength at the preceding visit (N.D., *Ct ≥* 43, 43 > *Ct ≥* 40, and 40 > *Ct*). Along with the first detection of *any* IS481 signal (model A: *C* ≤ 45), we also construct survival models predicting the time to first occurrence of intermediate and stronger IS481 signals (model B: *Ct* ≤ 43 and model C: *Ct* ≤ 40, respectively).

As we move from model A to model C, we focus on greater outcome specificity at the expense of fewer events and lower sensitivity. Overall, we find stronger evidence that infants (ego) are affected by mothers (alter) than vice versa. Figure 4 shows the hazard ratio (HR, Y axis) of each event (color) conditioned on the alter’s preceding visit (X axis), where the HR is relative to an N.D. in the alter’s preceding visit. Notably, we find that weaker IS481 signals in mothers’ preceding visit still predict elevated HRs in infants: *Ct* ≤ 45, HR=1.9 (1.3-2.8 95%CI); *Ct* ≤ 43, HR=2.0 (1.2-3.3 95%CI); *Ct* ≤ 40, HR=2.5 (0.98-6.1 95%CI). And, despite wide confidence intervals from infrequent events, a greatly increased HR of *Ct* ≤ 40 in both infants and mothers is predicted from preceding intermediate IS481 signals in their alters: infants, HR=6.3 (2.6-14.7 95%CI); mothers, HR=5.4 (1.9-14.2 95%CI).

**Figure 4:**
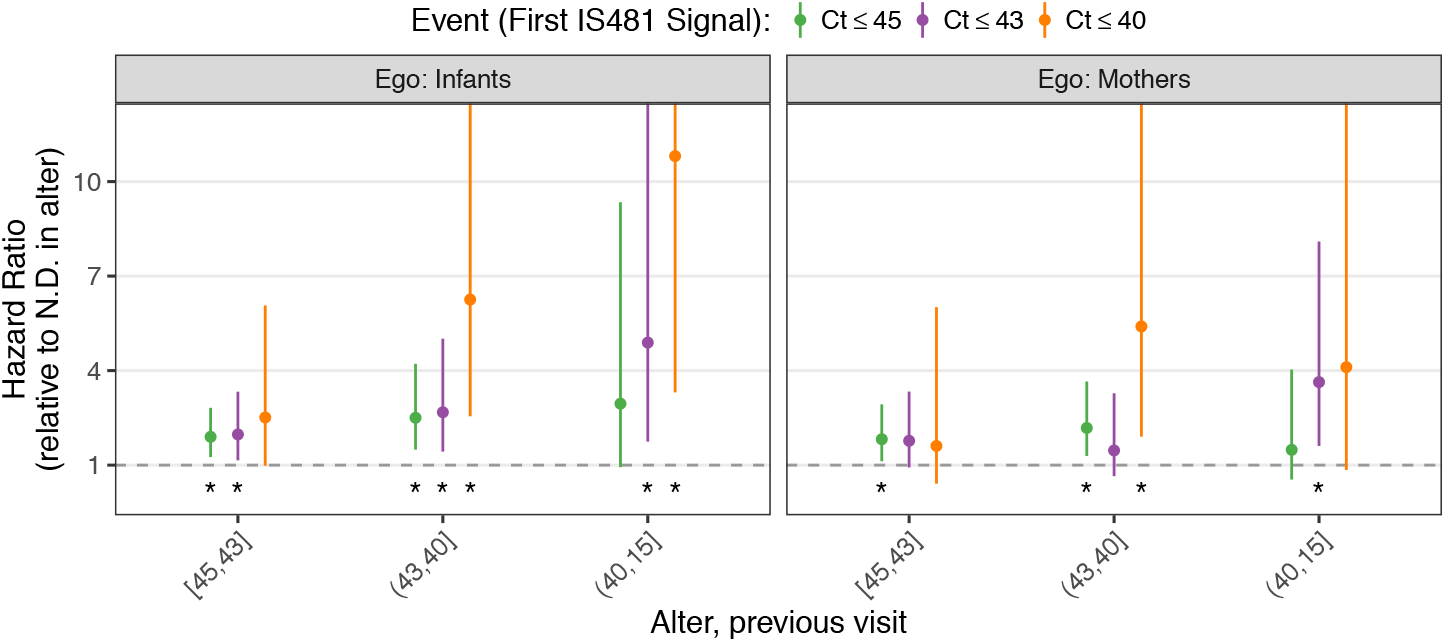
Survival analysis, showing hazard ratios (HR) of each event (color) in an ego conditioned on the alter’s IS481 signal in the previous visit (X axis; HR relative to N.D. in alter). Overall, HRs are higher in infants than mothers and are significantly greater than 1 in most cases (asterisk indicates 95% CI non-overlap with 1). Here ‘event’ denotes the first occurrence of an IS481 signal at least as strong as indicated. Infrequent events for the strongest signals yield large CI (*Ct* ≤ 40, upper limit not shown).

These results are broadly consistent with the assumption that mothers more frequently transmit pertussis to their infants than vice versa. Somewhat surprisingly, even weak signals in mothers correspond to elevated HRs in infants. And, while our study design can’t exclude the possibility of a shared source of infection, the time-ordered nature of these results offers a potentially useful clinical indicator for infant risk.

### Statistical associations within subjects and dyads

While the above survival models suggest a causal role of mothers’ IS481 signals on infants and (to a lesser extent) vice versa, these models only consider the first occurrence of each event. We next sought to characterize the full time course of each subject by considering two binary experimental outcomes: 1) future detections **within** subjects, and 2) contemporaneous detections within each dyad visit. We report the relative risk (RR) of each outcome stratified by IS481 signal strength, using N.D. as the baseline group.

For outcome 1, we predict future detections from each subject’s *own* IS481 signals at the preceding visit. For infants (Table 3a), we find an increase in the RR of detection from 2.3 (95% CI: 1.9-3.1) for weaker IS481 signals (*Ct ≥* 43) to 4.9 (95% CI: 3.7-6.5) for stronger signals (*Ct* < 40), and very little difference between infants and mothers (Table 3b).

**Table 3:** Relative risk of detection (RR +95%CI) per visit is greater given stronger IS481 *Ct* values in previous visits (no significant difference between (a) mothers and (b) infants).

| (a) |  |  |  | (b) |  |  |  |
| --- | --- | --- | --- | --- | --- | --- | --- |
| Infant IS481 | N.D. | Detect | RR | Mother IS481 | N.D. | Detect | RR |
| N.D. | 6240 | 525 | 1.0 | N.D. | 6159 | 527 | 1.0 |
| [45,43] | 263 | 62 | 2.5 (1.9-3.1) | [45,43] | 338 | 76 | 2.3 (1.9-2.9) |
| (43,40] | 159 | 55 | 3.3 (2.6-4.2) | (43,40] | 174 | 62 | 3.3 (2.7-4.2) |
| (40,15] | 53 | 33 | 4.9 (3.7-6.5) | (40,15] | 33 | 19 | 4.6 (3.2-6.7) |

For outcome 2, we predict detections in the ego from contemporaneous IS481 signals in their alter. We find a similar pattern as in outcome 1, where IS481 signals of mothers (alter) predict detections in infants (ego) and vice versa. Yet here we find notable differences between infants and mothers. For infants (Table 4a), we again find the RR of detection increasing from 2.7 (95% CI: 2.2-3.3) for weaker alter IS481 signals (*Ct ≥* 43) to 8.1 (95% CI: 6.3-10.4) for stronger signals (*Ct* < 40). We observe a more modest impact of infant *Ct* values on detections in mothers (Table 4b).

**Table 4:** Relative risk of contemporaneous detection (RR +95%CI) in ego given IS481 signal strength in alter: a) infant risk given mother’s IS481; b) mother’s risk given infant’s IS481. Overall, stronger signals predict more frequent detections, with modest differences between the predictive strength of IS481 signals in mothers a) versus infants b).

| (a) |  |  |  | (b) |  |  |  |
| --- | --- | --- | --- | --- | --- | --- | --- |
| Mother IS481 | N.D. | Detect | RR | Infant IS481 | N.D. | Detect | RR |
| N.D. | 7344 | 520 | 1.0 | N.D. | 7344 | 597 | 1.0 |
| [45,43] | 392 | 85 | 2.7 (2.2-3.3) | [45,43] | 293 | 91 | 3.2 (2.6-3.8) |
| (43,40] | 177 | 92 | 5.2 (4.3-6.2) | (43,40] | 166 | 79 | 4.3 (3.5-5.2) |
| (40,15] | 28 | 32 | 8.1 (6.3-10.4) | (40,15] | 61 | 39 | 5.2 (4.0-6.7) |

### Respiratory symptoms

Our final analysis of the full cohort investigates IS481 signal strength as a predictor of attendant respiratory symptoms. As above, we report RR for mothers and infants separately for *any* respiratory symptoms (predominantly cough and/or coryza). For infants, we also employ a set of GAMs to estimate the frequency of symptoms as a (continuous) response to IS481 signal strength. Here we focus on visits with IS481 detections (i.e., excluding N.D.s) and construct separate models for a) *any* respiratory symptoms and b) serious symptoms (i.e., excluding cough and/or coryza).

Unsurprisingly, we recorded respiratory symptoms far more frequently in infants than in mothers, regardless of IS481 signal strength (Table 5). Overall, we find that IS481 signals strongly predict symptoms in infants but only weakly in mothers. For infants (Table 5a), we see an increase in the RR of symptoms from 1.5 (95% CI: 1.2-1.9) for weaker IS481 signals (*Ct ≥* 43) to 5.1 (95% CI: 4.3-6.1) for stronger signals (40 > *Ct*). For mothers, we observe a (marginally significant) RR of symptoms that increases from 1.6 (95% CI: 1.1-2.3) for weaker IS481 signals to 2.4 (95% CI: 1.1-5.1) for stronger signals.

**Table 5:**
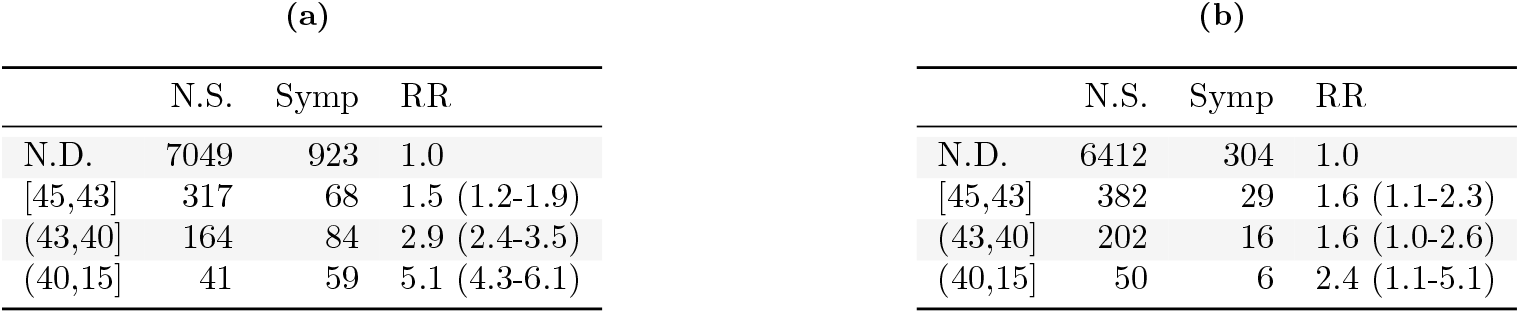
Relative risk of respiratory symptoms by IS481 signal strength for a) infants and b) mothers. No symptoms: N.S.

| (a) |  |  |  | (b) |  |  |  |
| --- | --- | --- | --- | --- | --- | --- | --- |
|  | N.S. | Symp | RR |  | N.S. | Symp | RR |
| N.D. | 7049 | 923 | 1.0 | N.D. | 6412 | 304 | 1.0 |
| [45,43] | 317 | 68 | 1.5 (1.2-1.9) | [45,43] | 382 | 29 | 1.6 (1.1-2.3) |
| (43,40] | 164 | 84 | 2.9 (2.4-3.5) | (43,40] | 202 | 16 | 1.6 (1.0-2.6) |
| (40,15] | 41 | 59 | 5.1 (4.3-6.1) | (40,15] | 50 | 6 | 2.4 (1.1-5.1) |

Focusing on infant visits with detections, Figure 5 shows the quantitative response of symptom frequency to IS481 *Ct*. The frequency of *any* symptom increases from 13% (95% CI: 9-18%) to 46% (95% CI: 40-52%) for *Ct* values of 45 and 40, respectively. While serious symptoms are much less common overall, again we find an increase from 2.5% (95% CI: 1.3-4.7%, *Ct* = 45) to 9% (95% CI: 6-13%, *Ct* = 40). We note that comparable models for mothers show no statistically significant association between IS481 *Ct* and frequency of respiratory symptoms.

**Figure 5:**
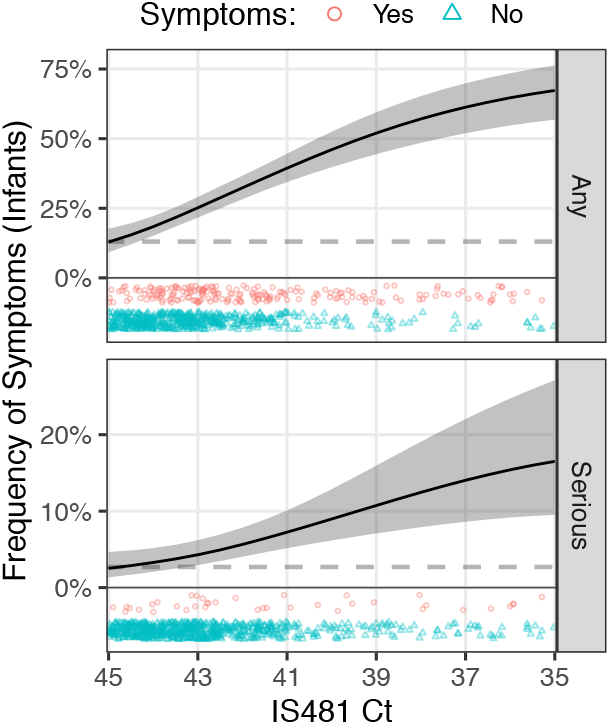
The frequency of reported respiratory symptoms increases with stronger IS481 signals among infant visits with IS481 detections (N=733), both for any symptoms (top) and serious symptoms (bottom: excludes solely cough and/or coryza). Curves show mean and 95% CI estimated from GAMs (one per row). Dashed lines shows marginal frequency of symptoms in infants (Any=13%; Serious=2.7%; N=8,705). Points show presence of respective symptoms by *Ct* value.

Taken together, these results quantify how IS481 signals cluster in time, within dyad visits, and correspond with respiratory symptoms. Notably, even the weakest IS481 signals (*Ct ≥* 43) are associated with statistically significant increases in the above outcomes (i.e., RR relative to N.D., Tables 3-5). We also observe noteworthy differences between infants and mothers, particularly for the time to first detection (Figure 4) and respiratory symptoms (Table 5 and Figure 5).

### Persistent colonization: evidence from a subgroup of subjects

The above analyses suggest an important role of false negatives in interpreting the time-course of individual subjects. Yet this potential ambiguity of qPCR signals complicates subject-level interpretation of infection status over time. Throughout the preceding analyses we have examined the full cohort, where IS481 signals are predominantly mild and infrequent (e.g., Fig 1C). To conclude, we examine those subjects with strong and/or frequent qPCR signals. In this way we focus on the time-course of subjects with *strong evidence of infection* (10), thereby reducing the relative impact of false negative qPCR tests.

We begin with a detailed view of 20 subjects (10 infants and 10 mothers) with the strongest *peak* IS481 signals, shown in Figure 6 (subjects ordered by number of detections). Clustering of detections around strong IS481 signals (*Ct* < 35) is evident here, consistent with our previous findings in this cohort of stepwise transitions of IS481 signals within subjects (i.e., Figure 6 in Gill and Gunning et al. (10)). However, uninterrupted runs of detections are uncommon. We also note that respiratory symptoms are common but not consistently observed in this group of subjects.

**Figure 6:**
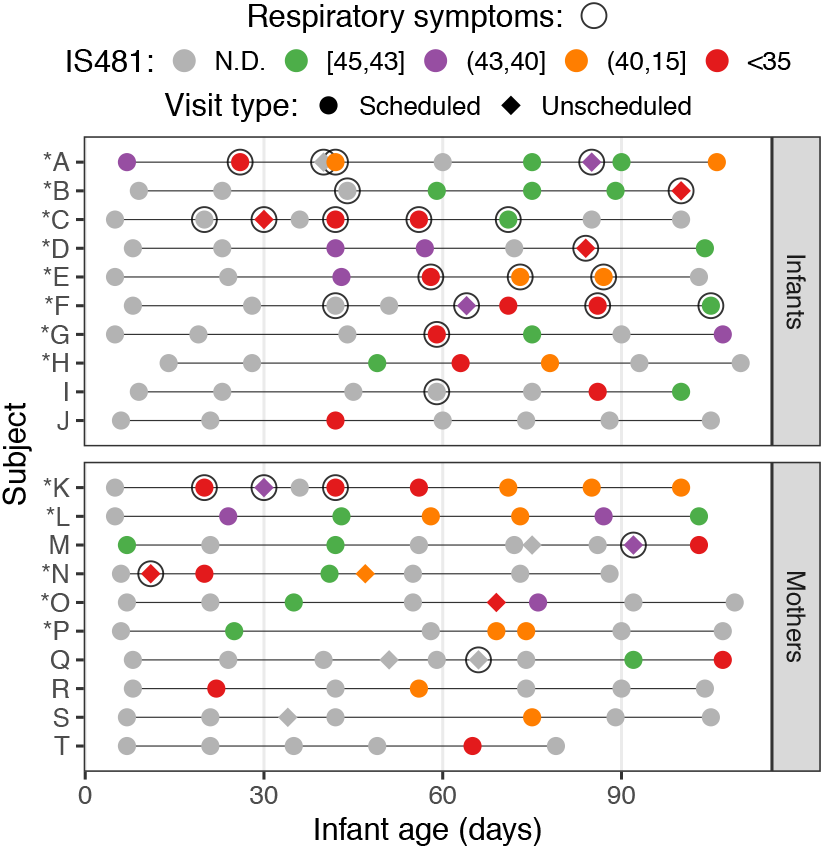
Timelines of study visits showing IS481 signals (color) for the ten infants (A-J) and ten mothers (K-T) with the strongest peak IS481 signals within each group. Peak IS481 *Ct* in these infants ranges from 18.1 to 32.6 (mean=27.1) and from 30.0 to 36.4 in these mothers (mean=33.7). Clusters of detections are evident that, however, frequently contain N.D.s. Within each panel, subjects are ordered by detection frequency. Open circles mark visits with any reported respiratory symptoms. Asterisks indicate inclusion in the subgroup analysis below (Fig 7), including 8 of 10 infants and 5 of 10 mothers.

With the above in mind, we sought to identify subjects with evidence of *persistent colonization*, which we define here as a cluster of *≥* 3 detections that lacks two or more adjacent N.D. (that is, we permit isolated but not consecutive N.D.s). In this way, we seek to balance sensitivity (including subjects with persistent evidence of IS481) with specificity (focusing on subjects where detections are clustered in time). Using these criteria, we identified a subgroup of 50 infants and 54 mothers. Despite not accounting for IS481 signal strength, this subgroup includes 8 infants and 5 mothers from Figure 6 (marked with asterisks above), showing that our duration-based criteria also capture many subjects of those exhibiting the strongest IS481 signals.

We provide a detailed analysis of this subgroup in Figure 7. Figure 7A displays the Kaplan-Meier estimated duration of detection, where no difference between infants and mothers is evident (median duration of 55 days, 95% CI: 46-69 (infants) and 50-68 days (mothers)). Figure 7B provides a detailed view of censoring, which was especially common in these mothers (where both left and right censoring occurred). Notably, the precision of the above duration estimates are limited by frequent right censoring (i.e., detection at last visit), which was observed in 20 infants and 20 mothers in the subgroup. Regardless, we find substantial evidence of persistent colonization, including 5 infants and 7 mothers with detections lasting *≥* 10 weeks (with right censoring in 5 of each).

**Figure 7:**
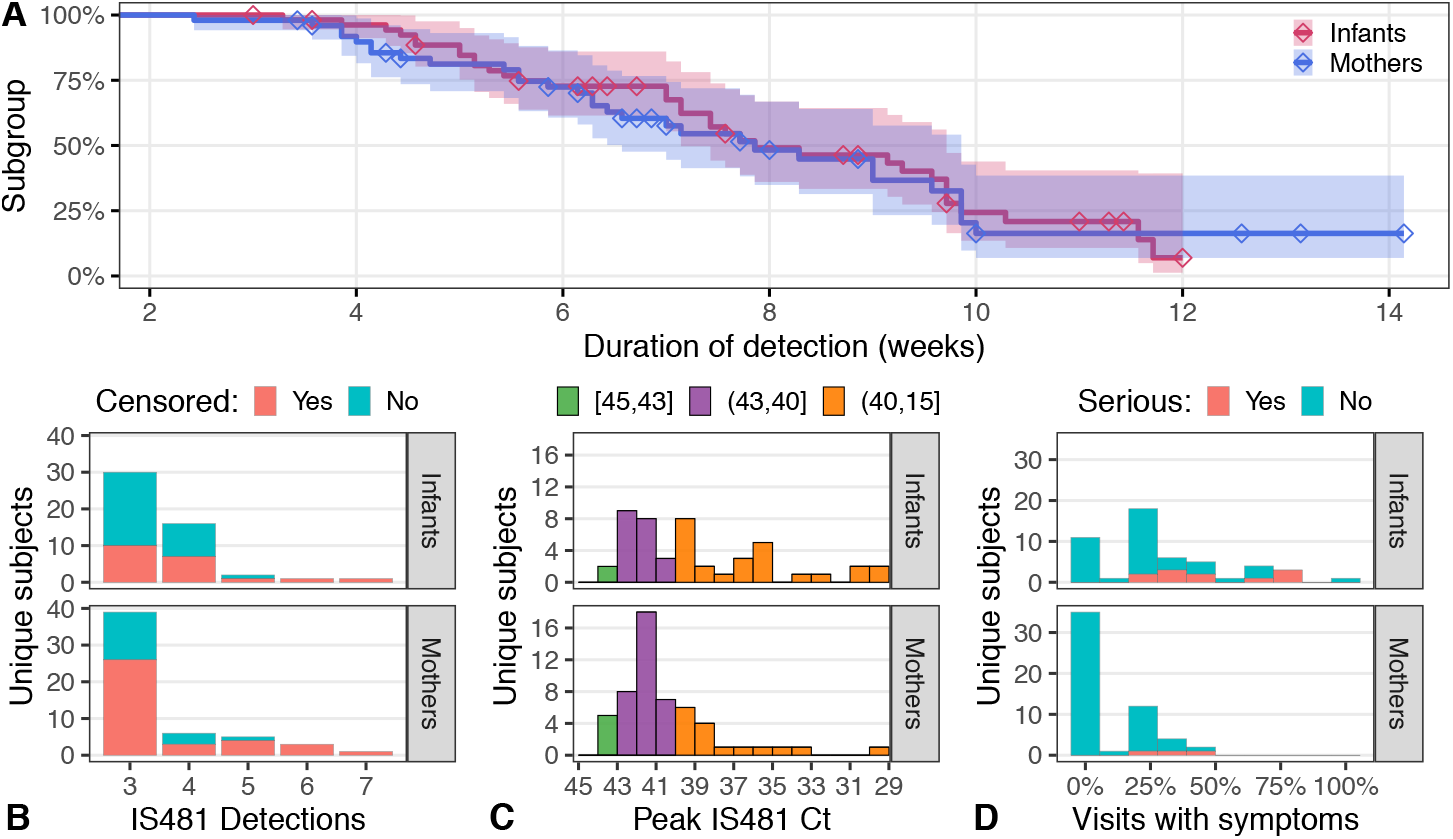
Subgroup analysis: subjects with evidence of persistent colonization, including 50 infants and 54 mothers. A) Kaplan-Meyer survival analysis of duration of detection (i.e., time interval between the first and last detection) that accounts for right censoring (detection at final study visit, shown by diamonds; includes 20 infants and 20 mothers). Estimated median duration of detection was 55 days. Panels B-D show the number of subjects by (B) number of detections, (C) peak IS481 signal, and (D) frequency of visits with respiratory symptoms. B) Number of detections stratified by censoring (color). For mothers, both left and right censoring are included (i.e., detection at first and last visit). C) Peak IS481 *Ct* is much higher in infants than mothers and, unlike the full cohort (Figure 1B), is rarely a weaker signal (green: *Ct* >43). Not shown: 3 infants with peak *Ct* ranging from 23.4 to 18.1. D) Most subgroup infants experienced respiratory symptoms infrequently, while most subgroup mothers experienced no respiratory symptoms. Color shows serious symptoms (i.e., excluding simple cough and/or coryza), which occurred most frequently in a small number of infants.

In Figures 7C-D, we illustrate peak IS481 *Ct* and frequency of respiratory symptoms. Figure 7C shows that peak IS481 *Ct* values are *much* higher in this subgroup than the full cohort (i.e., Figure 1B), and higher here in infants than mothers. Figure 7D highlights the relative sparsity of respiratory symptoms in this subgroup. We were particularly surprised to find that 22% of infants (N=11) and 65% of mothers (N=35) reported no symptoms across their study participation. In addition, of the 28 infants and 16 mothers with strong IS481 signals (Ct<40, Figure 7C), only 9 infants (32%) and 1 mother (6%) reported *any* serious respiratory symptoms.

Overall, we find strong evidence of persistent colonization in this subgroup, including strong peak IS481 signals and long durations of detection. In addition, frequent censoring shows that our 100-day follow-up period was not long enough to fully characterize the *maximum* duration of colonization, as evidenced by Figure 7A and 7C. Yet we observe minimal respiratory symptoms among this subgroup, even among infants. In addition, we find that a majority of these mothers reported no symptoms at all.

## Discussion

Pertussis remains a leading vaccine-preventable disease that is endemic worldwide, and whose global impacts have been underscored by its dramatic post-pandemic resurgence across a range of countries (48, 49, 50, 51, 1). This is despite generally high national vaccine coverage (44, 1), though considerable variation in vaccine uptake and timing exists worldwide (52, 53, 54, 41, 1). Further complicating this picture are major gaps in pertussis surveillance at the national and sub-national level, particularly in LMICs, as well as rapidly shifting diagnostic methods and standards (36, 55, 56, 57).

Meanwhile, mounting evidence of vaccine-driven evolution in *B. pertussis*, along with widespread antibiotic resistance in some regions, calls into question whether current clinical and public health tools will remain effective in the near future (58, 59, 26).

Within this milieu, sub-Saharan Africa represents a particular challenge, where poorly-resourced primary health care intersects with minimal surveillance and high disease burdens. For example, recent work revealed an almost complete lack of genomic surveillance within Africa, limiting conclusions about important aspects of pertussis epidemiology such as evolution of vaccine or antibiotics resistance (26, 57). Meanwhile, high fertility rates and HIV positivity throughout the region add to the supply of human hosts for disease transmission (60, 61, 62).

To successfully address the many public health challenges posed by pertussis, our results indicate that a clearer view of national and sub-national pertussis dynamics is needed, especially in low-resource settings where the burden of disease is highest. For example, our work points to the possibility of replacing symptoms-based pertussis surveillance with population-level qPCR-based prospective surveillance, which could then inform clinical care. In these regards, Lusaka is broadly representative of the urbanized Global South, where routine childhood vaccination coverage is generally high but community surveillance and clinical testing for pertussis is largely absent and prenatal maternal vaccination is cost-prohibitive (63).

In our cohort of mothers and their newborn infants, we rarely observed severe disease (see also Gill et al. (40)) but were surprised by the high overall frequency of IS481 detections, including a substantial number of intermediate and stronger IS481 signals (Figure 1). We commonly observed contemporaneous detections within mother-infant pairs (Table 1b), and we documented more frequent respiratory symptoms in those infants (but not mothers) with stronger IS481 signals (Table 5, Figure 5). We could not confidently attribute directionality of infection within dyads, nor can we definitively rule out shared environmental exposure. However, our survival analysis points to a greater impact of mothers on their infants than vice versa (Figure 4), a directionality that suggests causality.

Finally, we identified a subgroup of some 50 mothers and infants who displayed evidence of persistent colonization (median duration of approximately eight weeks), in which most mothers were entirely asymptomatic (Figure 7D). Taken together, our results support a picture of episodic but widespread pertussis transmission within the broader Chawama community that includes transmission from mostly asymptomatic mothers to their young infants.

Throughout this work, we have focused on on mild and minimally symptomatic pertussis, as classical pertussis was rare in our cohort (40). As noted above, we do not directly distinguish carriage from transmissible infection here, nor do we establish direct epidemiological linkage between cases. However, we can confidently state that A) evidence of infection (here, stronger IS481 signals) was much less common in mothers than in infants (Figure 1A-B) and that B) infants who experienced these stronger maternal IS481 signals were at greater risk of later exhibiting evidence of infection themselves (Figure 4). Furthermore, while both mild and more serious respiratory symptoms were commonly observed in infants with stronger IS481 signals, this pattern was largely absent for mothers (Table 5b and Figure 5 and 7). Overall, our results suggest that transmission from mothers to infants remains uncommon but that such transmission can nonetheless occur in the absence of symptomatic infection.

### Test reliability, disease surveillance, and public health

In this study, we exclusively employed qPCR-based testing due to its high sensitivity, relatively low cost, and minimal burden on study participants. In order to better interpret individual test results, we leveraged the temporal context of our longitudinal sampling scheme. With this approach, we considered all qPCR detections as potentially informative, allowing us to directly interrogate the predictive power of weaker versus stronger qPCR signals. As such, we do not directly distinguish carriage from transmissible infection (apart from statistical evidence, see Figure 4). We note, however, that the detailed relationship between exposure, carriage, and transmissible infection remains poorly characterized even when serological surveys are available (38, 39).

Considerable debate exists within the pertussis literature about the use of specific *Ct* thresholds for interpreting IS481 qPCR results in both clinical and epidemiological contexts (64, 65, 66, 15, 56). Our retesting results (Table 2 and Figure 3) emphasize the overall high fidelity of the IS481 qPCR assays conducted in our lab while also highlighting the sampling variation intrinsic in this assay (67, 65). In modern laboratory settings, this uncertainty is commonly addressed via repeat testing and qPCR efficiency correction (45, 47, 46). However, disease surveillance efforts are often time- and resource-constrained, especially in low-resource settings such as Lusaka. With this in mind, our central goal was *not* to directly quantify IS481 copy number, but rather to reliably identify and characterize pertussis infections in subjects. In this context, our limited qPCR retesting provides a benchmark for repeatability in this system and points to an important *Ct* threshold (*Ct* = 40) below which stochastic non-detections were rare in our lab.

In assessing the information content of weaker qPCR signals, some caveats are warranted. First and foremost, caution should be exercised in comparing *Ct* values between labs in the absence of objective standards and/or qPCR efficiency corrections (46, 68). We suspect that our average qPCR efficiency was lower than that of better-resourced laboratories, which would yield larger Ct values for a given sample. Notably, lower qPCR efficiency could partly explain the abundance of very weak IS481 signals in our results. Unfortunately, we did not retain PCR efficiency information as part of our laboratory pipeline. We recommend that future efforts prioritize reporting this information in accordance with modern best practices for Minimum Information for Publication of Quantitative Real-Time PCR Experiments (MIQE) guidelines (45).

We also note that interpretation of weak IS481 qPCR signals must account for the high copy number of this repetitive element. For example, our laboratory protocols specified that 1/50 of the DNA extracted from an NP sample was included in each qPCR test. Given a conservative estimate of 240 IS481 copies per genome, an NP sample that captured 10 bacteria would yield approximately 48 copies per qPCR test, well above the so-called qPCR limit of quantification of ten targets per reaction (see Fig. 8 in Ruiz-Villalba et al. (47)).

The above estimates stand in sharp contrast to the widely-cited claims of Tatti et al. (69), who write: *“On the basis of our analytical sensitivity data* … *IS481 CT values in the range of 35*<*CT*<*40 indicate the presence of less than 1 bacterium per reaction, which we consider uninterpretable*.*”* As we have detailed above, *Ct* values that represent fewer than one genome equivalent contain valuable information and should be expected in carefully-conducted *B. pertussis*surveillance efforts. We recognize that such weak and potentially ambiguous signals may not be appropriate for clinical diagnosis. However, our results demonstrate that they nonetheless contain valuable information about pathogen presence and infection intensity that can (and should) be leveraged for disease surveillance.

### Assessing population immunity (and susceptibility)

An important outstanding question from our study is the role of repeated pertussis infections in immune boosting. If we assume that 2015 was a typical year in Lusaka and that every mother who experienced *≥* 3 detections experienced a clinical infection (Figure 1C), then we estimate an annual attack rate of 6.8% of mothers. This corresponds to a mean return frequency of one infection per 14.8 years, which is much shorter than the presumed duration of immunity from natural infection or the whole-cell pertussis vaccination used in Zambia (70, 71). We note, however, ongoing debate surrounding the relative quality (and duration) of protection provided by natural infection, whole-cell vaccination and, more recently, a range of acellular vaccine formulations (72, 73, 74, 4, 1). We also note that pregnancy impacts immune functioning (75), such that the postpartum mothers in our cohort are not necessarily representative of the broader Chawama population. Nonetheless, our results point to episodically high community prevalence that could, in turn, drive immune boosting this population.

Unfortunately, the absence of unambiguous serological correlates of protection makes the confident assessment of population immunity via serosurveys challenging in the best of circumstances (76, 77, 39). And, while serological surveillance has been successfully deployed in similar settings (37), in our own field experiences, we found it much easier to increase the cohort size and frequency of clinic visits in the absence of blood draws and processing. With this in mind, we suggest that qPCR-based prospective disease surveillance offers a powerful and cost-efficient tool, both for ongoing pertussis surveillance and intervention activities as well as long-term research into pertussis transmission dynamics.

### Implications for global public health

Pertussis vaccine uptake remains a key country-level metric in tracking infectious disease prevention worldwide, led by the WHO and GAVI (78, 54, 79). However, much less is known about the true burden of pertussis infections in LMICs (63, 80, 81). In this context, pertussis surveillance in Lusaka is broadly representative of urban low-resource settings worldwide, where active surveillance is largely absent and passive surveillance only captures the most severe symptomatic illness. Indeed, the majority of active pertussis surveillance (and research) at present is focused on high-income countries where incidence is least common. In order to fully understand the “pertussis enigma” of sustained disease transmission in the face of high levels of presumed immunity from infection and/or vaccination, we expect that researchers will need to alter existing strategies and broaden their horizons.

As a populous, middle-income and primarily urban country, Zambia offers an evocative example of pertussis surveillance, where no cases have appeared in official WHO reports since 2009 (43). Yet by following a modest-sized cohort of 1,315 mothers and their newborn infants across a single year, we identified 55 infants (4.2%) and 50 mothers (3.8%) with evidence of persistent colonization, most of whom were minimally symptomatic. Taken together, these findings reaffirm the overall success of Zambia’s whole-cell pertussis immunization schedule in protecting against symptomatic disease. Moreover, our results are consistent with broad-based reductions in pertussis transmission at the population level. Yet these findings underscore both the stubborn persistence of pertussis within Zambia along with shortcomings of symptoms-based pertussis surveillance.

Our findings in Lusaka’s Chawama compound broadly align with the few available African studies of pertussis, including Uganda (15-20% prevalence in children with persistent coughs)(82) and South Africa’s notable PHIRST study (5.8-9.6% prevalence, mostly asymptomatic, in a prospective longitudinal household cohort) (56, 37) (see also Soofie et al. (83)). Overall, our results point to a long-overlooked “invisible mass” of minimally symptomatic pertussis incidence below the visible iceberg of officially recorded pertussis cases (84).

We believe our study also offers a template whereby high-quality disease surveillance can be achieved through efficient use of in-country public health resources. At various points we faced challenges in local capacity building and the timely processing of samples. Nonetheless, we believe that our strategy could be implemented at scale in a cost- and resource-efficient manner in LMICs, for example, through coordination of public health clinics with a central diagnostic laboratory, as we did. Such an active surveillance program would shed invaluable light on the total burden of pertussis incidence in low-resource settings, inform clinical treatment, and ultimately illuminate the factors that drive sustained transmission. Such a program could also complement local public health efforts, for example, by calibrating ongoing pertussis intervention efforts, and could yield further dividends by increasing public trust in government and building local biomedical diagnostic capacity (85).

## Materials and Methods

### Study Design

We provide a detailed account of study methods, including sample size considerations and inclusion criteria, in Gill et al. (40) and Gill et al. (10). We offer a brief overview here.

SAMIPS was designed as a longitudinal cohort study that followed mother/infant dyads across infants’ first three months of life. The study was based in Chawama compound, a densely populated low-resource peri-urban community near central Lusaka, Zambia. SAMIPS sought to enroll all healthy live births that occurred between March and December 2015 in Chawama, and included a public outreach campaign towards pregnant Chawama residents. Subjects were enrolled at the Chawama Primary Health Clinic (PHC), which is the only government-supported clinic in this community and primary source of medical care for Chawama residents. Mother/infant dyads were recruited at their first scheduled postpartum well-child visit (approximately 1 week of age). SAMIPS incentivized mothers to join and remain in the cohort by A) provided concierge (no-wait) routine and acute medical care for participants while enrolled, B) providing mothers with a per-visit travel stipend valued at approximately 7 US dollars and C), providing a small gift of baby supplies at the final scheduled study visit.

Each subject was scheduled to attend seven routine study visits, included a baseline enrollment visit and six follow-up visits spaced at 2–3 week intervals through 14 weeks old (maximum, 18 weeks). Additional unscheduled clinic visits could be initiated by mothers for acute medical care and/or routine well-child care. At each visit, we A) obtained nasopharyngeal (NP) swab samples from both mother and infant, B) recorded self-reported respiratory symptoms a standardized reporting sheet, and C) record administration of infant vaccinations and antibiotics (if any). To avoid possible contamination, routine childhood vaccinations were provided in a separate area of the clinic compound after NP sampling was conducted. We used Unique barcodes from pre-printed sticker books to track study records, thereby linking subjects, clinic visit records, and NP samples. qPCR results were unavailable during the study and thus could not affect clinical treatment.

The SAMIPS study received ethical approval from the Boston University School of Medicine IRB and the ERES Converge IRB in Lusaka. The data sets used in this analysis went through a de-identification process at the request of NIH, and no personally identifiable information is used here.

### Sample Collection and Processing

We collected NP samples using flocked-tipped nylon swabs (Copan Diagnostics, Merrieta, California). NP samples were collected from the PHC clinic daily and transported to our PCR laboratory at the University Teaching Hospital (UTH) and were stored at -80°C. DNA was extracted using the NucliSENS EasyMag system (bioMérieux, Marcy l’Etoile, France). Extracted DNA from each NP sample was tested for the IS481 insertion sequence and the constitutively expressed human RNase P (RNP) to assess sample collection, storage, DNA extraction, and qPCR reaction. Each 96-well qPCR plate contained a negative and IS481-positive control, along with approximately 46 samples (one each of IS481 and RNP). Reactions were run for 45 cycles on either a ABI 7500 thermocycler (ThermoFisher Scientific Inc, Waltham, MA) or a QuantStudio5 thermocycler (ThermoFisher Scientific Inc, Waltham, MA). We inspected samples run in parallel on both machines and found minimal systematic variation between machines. All primers and probes were purchased from Life Sciences Solutions (a subsidiary of ThermoFisher Scientific Inc).

For sample retests, a random subset of NP samples was drawn from the analysis set sample library stratified by IS481: we selected NP samples from 10 mothers and 10 infants across 5 evenly-spaced IS481 *Ct* intervals from *Ct* = 45 to *Ct* = 40, plus an additional interval of 40 > *Ct*.

### Statistical Analysis

Similar to our previous work (10), we focus here on dyads where both subjects had *≥* 4 fully-documented study visits. We exclude 6 mothers and 6 infants from our previous analysis set (i.e., Table 1 in Gill et al. (10)) where sufficient records for the full dyad was not available. All analysis was conducted in R 4.2.2 (86).

As in previous work, we define IS481 *detection* as any detectable *Ct* value (*Ct* > 45). We further categorized IS481 *Ct* values into *IS481 signal strength*: “no detection” (N.D.), weaker (*Ct* > 43), intermediate (43 > *Ct* > 40) and stronger (40 > *Ct*). These divisions were based on manual inspection of the empirical distribution of *Ct* values (Figure 1A) and model results from retest data (Figure 3B). Of note, we employ categorical IS481 signal strengths in order to compute relative risks for various quantities using N.D. as the reference level.

We also construct a set of linear models that employ IS481 *Ct* values as continuous predictors. We exclude N.D.s from these models, i.e., conditioning on detection. Generalized additive models (GAM) were constructed using the mgcv package (87). Confidence intervals for all linear models were computed via estimated marginal means using the emmeans package (88).

We constructed a three separate survival models for our (within subject) time-to-event analyses using a Weibull distribution. For each signal strength category, we predict the time (in days) to the first detection of at least that signal strength. Predictors include whether the subject is an infant or a mother and the alter’s categorical IS481 signal strength at the previous visit (and their interaction). We also tested models that included alters’ respiratory symptoms. Survival models and hazard ratios were fit using the flexsurv package (89). Hazard ratio confidence intervals were constructed via bootstrap simulations using 10,000 samples.

## Data Availability

Data available at https://osf.io/wya75/

https://osf.io/zjy4t

## Acknowledgements

This work would not have been possible without the gracious assistance of the Zambian Ministry of Health, the University Teaching Hospital in Lusaka, the Chawama Public Health Clinic, and Right to Care Zambia. In particular, we wish to thank Lawrence Mwananyanda, Geoffrey Kwenda, Zacharia Mupila, and Rachel C. Pieciak for their contributions to SAMIPS.

## Notes

### Competing Interest Statement

The authors have declared no competing interest.

### Funding Statement

This study was funded by the NIH (R01AI133080) and Bill and Melinda Gates Foundation (OPP1105094).

### Author Declarations

The data for this analysis came from the Southern African Mother Infant Pertussis study (SAMIPS), which received ethical approval from the ERES converge IRB in Lusaka, and the IRB at Boston University Medical Center. The current analysis uses a de-identified data set at the request of the NIH for the R01 grant that supported this work. The current data sets contains no personally identifiable information.

### Summary of Updates

Revisions to text based on comments from authors: primarily Intro and Discussion, along with additional literature cited. The layout of Fig 4 has been modified but underlying data are unchanged. See public reviews here: https://elifesciences.org/reviewed-preprints/106139/reviews#tab-content

## References

[1] Matthieu Domenech de Cellés and Pejman Rohani. Pertussis vaccines, epidemiology and evolution. Nature Reviews Microbiology, pages 1–14, 2024. ISSN 1740-1526. doi: 10.1038/s41579-024-01064-8.

[2] J D Cherry. The Science and Fiction of the “Resurgence” of Pertussis. Pediatrics, 112(2):405–406, 02 2003. doi: 10.1542/peds.112.2.405.

[3] M J Bart, S R Harris, A Advani, Y Arakawa, D Bottero, V Bouchez, P K Cassiday, C S Chiang, T Dalby, N K Fry, M E Gaillard, M Van Gent, N Guiso, H O Hallander, E T Harvill, Q He, H G J Van der Heide, K Heuvelman, D F Hozbor, K Kamachi, G I Karataev, R Lan, A Luty ska, R P Maharjan, J Mertsola, T Miyamura, S Octavia, A Preston, M A Quail, V Sintchenko, P Stefanelli, M L Tondella, R S W Tsang, Y Xu, S M Yao, S Zhang, J Parkhill, and F R Mooi. Global Population Structure and Evolution of Bordetella pertussis and Their Relationship with Vaccination. mBio, 5(2):e01074.–14–e01074–14, 02 2014. doi: 10.1128/mbio.01074-14.

[4] Pejman Rohani and Samuel V Scarpino. Pertussis: Epidemiology, Immunology & Evolution. Oxford Univ Press. Oxford Univ Press, 04 2019.

[5] Paul E Kilgore, Abdulbaset M Salim, Marcus J Zervos, and Heinz-Josef Schmitt. Pertussis: Microbiology, Disease, Treatment, and Prevention. Clinical Microbiology Reviews, 29(3):449–486, 03 2016. doi: 10.1128/cmr.00083-15.

[6] Tina Tan, Evelinda Trindade, and Danuta Skowronski. Epidemiology of pertussis. The Pediatric Infectious Disease Journal, 24(Supplement):S10–S18, 2005. doi: 10.1097/01.inf.0000160708.43944.99.

[7] Florens GA Versteegh, Joop FP Schellekens, André Fleer and John J Roord. Pertussis: a concise historical review including diagnosis, incidence, clinical manifestations and the role of treatment and vaccination in management. Reviews in Medical Microbiology, 16(3):79–89, 2005. ISSN 0954-139X. doi: 10.1097/01.revmedmi.0000175933.85861.4e.

[8] Kevin D Forsyth, Carl-Heinz Wirsing von Konig, Tina Tan, Jaime Caro, and Stanley Plotkin. Prevention of pertussis: recommendations derived from the second Global Pertussis Initiative roundtable meeting. Vaccine, 25(14):2634–2642, 03 2007. doi: 10.1016/j.vaccine.2006.12.017.

[9] Rodger Craig, Elizabeth Kunkel, Natasha S Crowcroft, Meagan C Fitzpatrick, Hester de Melker, Benjamin M Althouse, Tod Merkel, Samuel V Scarpino, Katia Koelle, Lindsay Friedman, Callum Arnold, and Shelly Bolotin. Asymptomatic Infection and Transmission of Pertussis in Households: A Systematic Review. Clinical infectious diseases, 70 (1):152–161, 01 2020. doi: 10.1093/cid/ciz531.

[10] Christopher J Gill, Christian E Gunning, William B MacLeod, Lawrence Mwananyanda, Donald M Thea, Rachel C Pieciak, Geoffrey Kwenda, Zacharia Mupila, and Pejman Rohani. Asymptomatic Bordetella pertussis infections in a longitudinal cohort of young African infants and their mothers. eLife, 10:e65663, 2021. doi: 10.7554/elife.65663.

[11] JD Cherry, LJ Baraff, and E Hewlett. The past, present, and future of pertussis. The role of adults in epidemiology and future control. Western journal of medicine, 150(3):319, 1989.

[12] C.H Wirsing von König, S Postels-Multan, H.J Schmitt, and H.L Bock. Pertussis in adults: frequency of transmission after household exposure. The Lancet, 346(8986):1326–1329, 1995. ISSN 0140-6736. doi: 10.1016/s0140-6736(95)92343-8.

[13] Joop Schellekens, Carl-Heinz Wirsing von König, and Pierce Gardner. Pertussis Sources of Infection and Routes of Transmission in the Vaccination Era. The Pediatric Infectious Disease Journal, 24(5):S19–S24, 2005. ISSN 0891-3668. doi: 10.1097/01.inf.0000160909.24879.e6.

[14] Hans de Graaf, Muktar Ibrahim, Alison R Hill, Diane Gbesemete, Andrew T Vaughan, Andrew Gorringe, Andrew Preston, Annemarie M Buisman, Saul N Faust, Kent E Kester, Guy A M Berbers, Dimitri A Diavatopoulos, and Robert C Read. Controlled Human Infection With Bordetella pertussis Induces Asymptomatic, Immunizing Colonization. Clinical Infectious Diseases: An Official Publication of the Infectious Diseases Society of America, 71(2): 403–411, 2020. ISSN 1058-4838. doi: 10.1093/cid/ciz840.

[15] Shelly Bolotin, Helen Quinn, and McIntyre, and Peter. Surveillance and diagnostics. In Rohani and Pejman, editors, Pertussis: Epidemiology, Immunology & Evolution, pages 193–210. Oxford University Press, 2018.

[16] Edward L Chan, Nick Antonishyn, Ryan McDonald, Tara Maksymiw, Peter Pieroni, Evelyn Nagle, and Greg B Horsman. The use of taqman pcr assay for detection of bordetella pertussis infection from clinical specimens. Archives of pathology & laboratory medicine, 126(2):173–176, 2002.

[17] Norman K Fry, Oceanis Tzivra, Y Ting Li, Anthony McNiff, Nivedita Doshi, PA Christopher Maple, Natasha S Crowcroft, Elizabeth Miller, Robert C George, and Timothy G Harrison. Laboratory diagnosis of pertussis infections: the role of pcr and serology. Journal of medical microbiology, 53(6):519–525, 2004.

[18] Alberto E Tozzi, Lucia Pastore Celentano, Marta Luisa Ciofi degli Atti, and Stefania Salmaso. Diagnosis and management of pertussis. CMAJ : Canadian Medical Association journal = journal de l’Association medicale canadienne, 172(4):509–515, 02 2005. doi: 10.1503/cmaj.1040766.

[19] PE Fine and JA Clarkson. Distribution of immunity to pertussis in the population of England and Wales. The Journal of Hygiene, 92(1):21–26, 1984.

[20] C.E. Gunning, E. Erhardt, and H.J. Wearing. Conserved patterns of incomplete reporting in pre-vaccine era childhood diseases. Proc. R. Soc. B, 281(1794):20140886, 2014.

[21] Christian E Gunning, Matthew J Ferrari, Erik B Erhardt, and Helen J Wearing. Evidence of cryptic incidence in childhood diseases. Proceedings of the Royal Society B: Biological Sciences, 284(1861):20171268, 2017.

[22] Pejman Rohani,J J Earn, and B T Grenfell. Opposite patterns of synchrony in sympatric disease metapopulations. Science, 286(5441):968–971, 10 1999.

[23] Willem G van Panhuis, John Grefenstette, Su Yon Jung, Nian Shong Chok, Anne Cross, Heather Eng, Bruce Y Lee, Vladimir Zadorozhny, Shawn Brown, Derek Cummings, and Donald S Burke. Contagious diseases in the United States from 1888 to the present. The New England Journal of Medicine, 369(22):2152–2158, 11 2013. doi: 10.1056/nejmms1215400.

[24] Maria Yui Kwan Chow, Gulam Khandaker, and Peter McIntyre. Global Childhood Deaths From Pertussis: A Historical Review. Clinical Infectious Diseases, 63(suppl 4):S134–S141, 2016. ISSN 1058-4838. doi: 10.1093/cid/ciw529.

[25] World Health Organization. Pertussis vaccines: WHO position paper. Weekly Epidemiological Record, 90:1–28, 09 2015.

[26] Noémie Lefrancq, Valérie Bouchez, Nadia Fernandes, Alex-Mikael Barkoff, Thijs Bosch, Tine Dalby, Thomas åkerlund, Jessica Darenberg, Katerina Fabianova, Didrik F. Vestrheim, Norman K. Fry, Juan José González-López, Karolina Gullsby, Adele Habington, Qiushui He, David Litt, Helena Martini, Denis Piérard, Paola Stefanelli, Marc Stegger, Jana Zavadilova, Nathalie Armatys, Annie Landier, Sophie Guillot, Samuel L. Hong, Philippe Lemey, Julian Parkhill, Julie Toubiana, Simon Cauchemez, Henrik Salje, and Sylvain Brisse. Global spatial dynamics and vaccine-induced fitness changes of Bordetella pertussis. Science Translational Medicine, 14(642):eabn3253, 2022. ISSN 1946-6234. doi: 10.1126/scitranslmed.abn3253.

[27] F R Mooi, N A T van der MAAS, and H E De Melker. Pertussis resurgence: waning immunity and pathogen adaptation – two sides of the same coin. Epidemiology and Infection, pages 1–10, 02 2013. doi: 10.1017/s0950268813000071.

[28] Julian Parkhill, Mohammed Sebaihia, Andrew Preston, Lee D Murphy, Nicholas Thomson, David E Harris, Matthew T G Holden, Carol M Churcher, Stephen D Bentley, Karen L Mungall, Ana M Cerdeño-Tárraga, Louise Temple, Keith James, Barbara Harris, Michael A Quail, Mark Achtman, Rebecca Atkin, Steven Baker, David Basham, Nathalie Bason, Inna Cherevach, Tracey Chillingworth, Matthew Collins, Anne Cronin, Paul Davis, Jonathan Doggett, Theresa Feltwell, Arlette Goble, Nancy Hamlin, Heidi Hauser, Simon Holroyd, Kay Jagels, Sampsa Leather, Sharon Moule, Halina Norberczak, Susan O’Neil, Doug Ormond, Claire Price, Ester Rabbinowitsch, Simon Rutter, Mandy Sanders, David Saunders, Katherine Seeger, Sarah Sharp, Mark Simmonds, Jason Skelton, Robert Squares, Steven Squares, Kim Stevens, Louise Unwin, Sally Whitehead, Bart G Barrell, and Duncan J Maskell. Comparative analysis of the genome sequences of Bordetella pertussis, Bordetella parapertussis and Bordetella bronchiseptica. nature genetics, 35(1):32–40, 08 2003. doi: 10.1126/science.7973728.

[29] Katie L Sealey, Thomas Belcher, and Andrew Preston. Bordetella pertussis epidemiology and evolution in the light of pertussis resurgence. MEEGID, 40:136–143, 06 2016. doi: 10.1016/j.meegid.2016.02.032.

[30] Michael R Weigand, Yanhui Peng, Vladimir Loparev, Dhwani Batra, Katherine E Bowden, Mark Burroughs, Pamela K Cassiday, Jamie K Davis, Taccara Johnson, Phalasy Juieng, Kristen Knipe, Marsenia H Mathis, Andrea M Pruitt, Lori Rowe, Mili Sheth, M Lucia Tondella, and Margaret M Williams. The History of Bordetella pertussis Genome Evolution Includes Structural Rearrangement. Journal of Bacteriology, 199(8), 2017. ISSN 0021-9193. doi: 10.1128/jb.00806-16.

[31] Lind-Brandberg, Welinder-Olsson, Lagergård, Taranger Trollfors, and & Zackrisson. Evaluation of PCR for diagnosis of Bordetella pertussis and Bordetella parapertussis infections. Journal of clinical microbiology, 36(3):679–683, 1998.

[32] Hans O Hallander. Microbiological and Serological Diagnosis of Pertussis. Clinical Infectious Diseases, 28(2):S99–S106, 1999. ISSN 1058-4838. doi: 10.1086/515056.

[33] C H Wirsing von König, S Halperin, M Riffelmann, and N Guiso. Pertussis of adults and infants. The Lancet Infectious Diseases, 2(12):744–750, 12 2002.

[34] Adria D Lee, Pamela K Cassiday, Lucia C Pawloski, Kathleen M Tatti, Monte D Martin, Elizabeth C Briere, M Lucia Tondella, Stacey W Martin, and Clinical Validation Study Group. Clinical evaluation and validation of laboratory methods for the diagnosis of bordetella pertussis infection: culture, polymerase chain reaction (pcr) and anti-pertussis toxin igg serology (igg-pt). PLoS One, 13(4):e0195979, 2018.

[35] Aaron Mark Wendelboe and Annelies Van Rie. Diagnosis of pertussis: a historical review and recent developments. Expert review of molecular diagnostics, 6(6):857–864, 2006.

[36] Anneke van der Zee, Joop FP Schellekens, and Frits R Mooi. Laboratory diagnosis of pertussis. Clinical microbiology reviews, 28(4):1005–1026, 2015.

[37] Fahima Moosa, Jackie Kleynhans, Lillian Makhathini, Mignon du Plessis, Stefano Tempia, Meredith L McMorrow, Jocelyn Moyes, Amelia Buys, Lorens Maake, Sheilagh Smit, et al. Bordetella pertussis infection and antibody dynamics in household cohorts in two south african communities, 2016–2018: findings from the phirst study. Journal of Infection, page 106550, 2025.

[38] Rodger Craig, Elizabeth Kunkel, Natasha S Crowcroft, Meagan C Fitzpatrick, Hester De Melker, Benjamin M Althouse, Tod Merkel, Samuel V Scarpino, Katia Koelle, Lindsay Friedman, et al. Asymptomatic infection and transmission of pertussis in households: a systematic review. Clinical infectious diseases, 70(1):152–161, 2020.

[39] Matthieu Domenech de Cellés, Anabelle Wong, Tine Dalby, and Pejman Rohani. Natural immune boosting biases pertussis infection estimates in seroprevalence studies. Nature Communications, 16(1):8883, 2025.

[40] Christopher J Gill, Lawrence Mwananyanda, William MacLeod, Geoffrey Kwenda, Magdalene Mwale, Anna L Williams, Kazungu Siazeele, Zhaoyan Yang, James Mwansa, and Donald M Thea. Incidence of severe and nonsevere pertussis among hiv-exposed and-unexposed zambian infants through 14 weeks of age: results from the southern africa mother infant pertussis study (samips), a longitudinal birth cohort study. Clinical Infectious Diseases, 63(suppl 4): S154–S164, 2016.

[41] Christian E Gunning, Lawrence Mwananyanda, William B MacLeod, Magdalene Mwale, Donald M Thea, Rachel C Pieciak, Pejman Rohani, and Christopher J Gill. Implementation and adherence of routine pertussis vaccination (dtp) in a low-resource urban birth cohort. BMJ open, 10(12):e041198, 2020.

[42] Christian E Gunning, Pejman Rohani, Lawrence Mwananyanda, Geoffrey Kwenda, Zacharia Mupila, and Christopher J Gill. Young zambian infants with symptomatic rsv and pertussis infections are frequently prescribed inappropriate antibiotics: a retrospective analysis. PeerJ, 11:e15175, 2023.

[43] World Health Organization. Pertussis reported cases and incidence. https://immunizationdata.who.int/global/wiise-detail-page/pertussis-reported-cases-and-incidence. Accessed 2024-10-11.

[44] World Health Organization. Diphtheria tetanus toxoid and pertussis (dtp) vaccination coverage. https://immunizationdata.who.int/global/wiise-detail-page/diphtheria-tetanus-toxoid-and-pertussis-(dtp)-vaccination-coverage?GROUP=WHO_REGIONS&ANTIGEN=&YEAR=&CODE=. [Accessed 2024-10-11].

[45] Stephen A Bustin, Vladimir Benes, Jeremy A Garson, Jan Hellemans, Jim Huggett, Mikael Kubista, Reinhold Mueller, Tania Nolan, Michael W Pfaffl, Gregory L Shipley, et al. The miqe guidelines: Minimum information for publication of quantitative real-time pcr experiments. Clinical Chemistry, 55(4):611–622, 2009.

[46] Jan M Ruijter, Rebecca J Barnewall, Ian B Marsh, Andrew N Szentirmay, Jane C Quinn, Robin van Houdt, Quinn D Gunst, and Maurice JB van den Hoff. Efficiency correction is required for accurate quantitative pcr analysis and reporting. Clinical chemistry, 67(6):829–842, 2021.

[47] Adrián Ruiz-Villalba, Jan M Ruijter, and Maurice JB van den Hoff. Use and misuse of cq in qpcr data analysis and reporting. Life, 11(6):496, 2021.

[48] Pejman Rohani and John M Drake. The decline and resurgence of pertussis in the US. Epidemics, 3(3–4):183–188, 2011. doi: 10.1016/j.epidem.2011.10.001.

[49] Matthieu Domenech de Cellés, Felicia M G Magpantay, Aaron A King, and Pejman Rohani. The pertussis enigma: reconciling epidemiology, immunology and evolution. Proceedings of the Royal Society B: Biological Sciences, 283 (1822):20152309, 01 2016. doi: 10.1098/rspb.2015.2309.

[50] Yoon Hong Choi, Helen Campbell, Gayatri Amirthalingam, Albert Jan Van Hoek, and Elizabeth Miller. Investigating the pertussis resurgence in England and Wales, and options for future control. BMC Medicine, 14:1–11, 08 2016. doi: 10.1186/s12916-016-0665-8.

[51] Patricia Therese Campbell, James Matthew McCaw, Peter McIntyre, and Jodie McVernon. Defining long-term drivers of pertussis resurgence, and optimal vaccine control strategies. Vaccine, 33(43):5794–5800, 09 2015. doi: 10.1016/j.vaccine.2015.09.025.

[52] Andrew Clark and Colin Sanderson. Timing of children’s vaccinations in 45 low-income and middle-income countries: an analysis of survey data. The Lancet, 373(9674):1543–1549, 2009.

[53] Manas K Akmatov and Rafael T Mikolajczyk. Timeliness of childhood vaccinations in 31 low and middle-income countries. J Epidemiol Community Health, 66(7):e14–e14, 2012.

[54] Jonathan F Mosser, William Gagne-Maynard, Puja C Rao, Aaron Osgood-Zimmerman, Nancy Fullman, Nicholas Graetz, Roy Burstein, Rachel L Updike, Patrick Y Liu, Sarah E Ray, et al. Mapping diphtheria-pertussis-tetanus vaccine coverage in africa, 2000–2016: a spatial and temporal modelling study. The Lancet, 393(10183):1843–1855, 2019.

[55] Amanda E Faulkner, Tami H Skoff, M Lucia Tondella, Amanda Cohn, Thomas A Clark, and Stacey W Martin. Trends in pertussis diagnostic testing in the united states, 1990 to 2012. The Pediatric infectious disease journal, 35 (1):39–44, 2016.

[56] Fahima Moosa, Mignon du Plessis, Nicole Wolter, Maimuna Carrim, Cheryl Cohen, Claire von Mollendorf, Sibongile Walaza, Stefano Tempia, Halima Dawood, Ebrahim Variava, et al. Challenges and clinical relevance of molecular detection of bordetella pertussis in south africa. BMC Infectious Diseases, 19:1–11, 2019.

[57] Fahima Moosa, Mignon du Plessis, Michael R Weigand, Yanhui Peng, Dineo Mogale, Linda de Gouveia, Marta C Nunes, Shabir A Madhi, Heather J Zar, Gary Reubenson, et al. Genomic characterization of bordetella pertussis in south africa, 2015–2019. Microbial Genomics, 9(12):001162, 2023.

[58] Alex-Mikael Barkoff, Jussi Mertsola, Denis Pierard, Tine Dalby, Silje Vermedal Hoegh, Sophie Guillot, Paola Stefanelli, Marjolein van Gent, Guy Berbers, Didrik F Vestrheim, Margrethe Greve-Isdahl, Lena Wehlin, Margaretha Ljungman, Norman K Fry, Kevin Markey, Kari Auranen, and Qiushui He. Surveillance of Circulating Bordetella pertussis Strains in Europe during 1998 to 2015. Journal of Clinical Microbiology, 56(5):ftv050, 2018. doi: 10.1016/j.vaccine.2009.07.010.

[59] Laurence Don Wai Luu, Raisa Rafique, Michael Payne, Sophie Octavia, Jennifer Robson, Vitali Sintchenko, and Ruiting Lan. Deciphering bordetella pertussis epidemiology through culture-independent multiplex amplicon and metagenomic sequencing. Journal of Clinical Microbiology, pages e01178–24, 2024.

[60] Desalegn Tesfa, Sofonyas Abebaw Tiruneh, Alemayehu Digssie Gebremariam, Melkalem Mamuye Azanaw, Melaku Tadege Engidaw, Belayneh Kefale, Bedilu Abebe, Tsion Dessalegn, and Mulu Tiruneh. The pooled estimate of the total fertility rate in sub-saharan africa using recent (2010–2018) demographic and health survey data. Frontiers in Public Health, 10:1053302, 2023.

[61] Jane N O’Sullivan. Demographic delusions: World population growth is exceeding most projections and jeopardising scenarios for sustainable futures. World, 4(3):545–568, 2023.

[62] Abdulhammed O Babatunde, Oluwawapelumi D Akin-Ajani, Ridwanullah O Abdullateef, Taofeeq O Togunwa, and Haroun O Isah. Review of antiretroviral therapy coverage in 10 highest burden hiv countries in africa: 2015–2020. Journal of Medical Virology, 95(1):e28320, 2023.

[63] Rudzani Muloiwa, Nicole Wolter, Ezekiel Mupere, Tina Tan, AJ Chitkara, Kevin D Forsyth, Carl-Heinz Wirsing von König, and Gregory Hussey. Pertussis in africa: findings and recommendations of the global pertussis initiative (gpi). Vaccine, 36(18):2385–2393, 2018.

[64] Valerie Waters, Frances Jamieson, Susan E Richardson, Michael Finkelstein, Anne Wormsbecker, and Scott A Halperin. What is the significance of a high cycle threshold positive is481 pcr for bordetella pertussis? The Pediatric infectious disease journal, 28(12):1143–1144, 2009.

[65] Kathleen M Tatti, Stacey W Martin, Kathryn O Boney, Kristin Brown, Thomas A Clark, and Maria Lucia Tondella. Qualitative assessment of pertussis diagnostics in united states laboratories. The Pediatric infectious disease journal, 32(9):942–945, 2013.

[66] Shelly Bolotin, Shelley L Deeks, Alex Marchand-Austin, Heather Rilkoff, Vica Dang, Ryan Walton, Ahmed Hashim, David Farrell, and Natasha S Crowcroft. Correlation of real time pcr cycle threshold cut-off with bordetella pertussis clinical severity. PLoS One, 10(7):e0133209, 2015.

[67] Kathleen M Tatti, Kai-Hui Wu, Maria Lucia Tondella, Pamela K Cassiday, Margaret M Cortese, Patricia P Wilkins, and Gary N Sanden. Development and evaluation of dual-target real-time polymerase chain reaction assays to detect bordetella spp. Diagnostic microbiology and infectious disease, 61(3):264–272, 2008.

[68] Andreas Untergasser, Jan M Ruijter, Vladimir Benes, and Maurice JB van den Hoff. Web-based linregpcr: application for the visualization and analysis of (rt)-qpcr amplification and melting data. BMC bioinformatics, 22(1):398, 2021.

[69] Kathleen M Tatti, Kansas N Sparks, Kathryn O Boney, and Maria Lucia Tondella. Novel multitarget real-time pcr assay for rapid detection of bordetella species in clinical specimens. Journal of clinical microbiology, 49(12):4059–4066, 2011.

[70] Helen J Wearing and Pejman Rohani. Estimating the Duration of Pertussis Immunity Using Epidemiological Signatures. PLoS Pathogens, 5(10):e1000647, 10 2009. doi: 10.1371/journal.ppat.1000647.s012.

[71] Manoj Gambhir, Thomas A. Clark, Simon Cauchemez, Sara Y. Tartof, David L. Swerdlow, and Neil M. Ferguson. A Change in Vaccine Efficacy and Duration of Protection Explains Recent Rises in Pertussis Incidence in the United States. PLoS Computational Biology, 11(4):e1004138, 2015. ISSN 1553-734X. doi: 10.1371/journal.pcbi.1004138.

[72] Paul EM Fine and JacquelineA Clarkson. The recurrence of whooping cough: possible implications for assessment of vaccine efficacy. The Lancet, 319(8273):666–669, 1982.

[73] Pejman Rohani, David JD Earn, and Bryan T Grenfell. Impact of immunisation on pertussis transmission in england and wales. The Lancet, 355(9200):285–286, 2000.

[74] James D Cherry. The epidemiology of pertussis: a comparison of the epidemiology of the disease pertussis with the epidemiology of bordetella pertussis infection. Pediatrics, 115(5):1422–1427, 2005.

[75] Michael Pazos, Rhoda S Sperling, Thomas M Moran, and Thomas A Kraus. The influence of pregnancy on systemic immunity. Immunologic research, 54:254–261, 2012.

[76] Kingston H.G. Mills. Immunity to Bordetella pertussis. Microbes and Infection, 3(8):655–677, 07 2001. ISSN 1286-4579. doi: 10.1016/s1286-4579(01)01421-6.

[77] Mieszko M. Wilk, Aideen C. Allen, Alicja Misiak, Lisa Borkner, and Kingston H.G. Mills. The immunology of Bordetella pertussis infection and vaccination. In Pertussis: Epidemiology, Immunology & Evolution. Oxford University Press, 2019.

[78] Jean-Marie Okwo-Bele and Thomas Cherian. The expanded programme on immunization: a lasting legacy of smallpox eradication. Vaccine, 29:D74–D79, 2011.

[79] Robert John Kolesar, Tsheten Tsheten, et al. Evaluating country performance after transitioning from gavi assistance: an applied synthetic control analysis. Global Health: Science and Practice, 11(4), 2023.

[80] Ajoke Sobanjo-ter Meulen, Philippe Duclos, Peter McIntyre, Kristen D. C. Lewis, Pierre Van Damme, Katherine L. O’Brien, and Keith P. Klugman. Assessing the evidence for maternal pertussis immunization: A report from the bill & melinda gates foundation symposium on pertussis infant disease burden in low- and lower-middle-income countries. Clinical Infectious Diseases, 63(Suppl 4):S123–S133, 2016.

[81] Rudzani Muloiwa, Benjamin M Kagina, Mark E Engel, and Gregory D Hussey. The burden of laboratory-confirmed pertussis in low-and middle-income countries since the inception of the expanded programme on immunisation (epi) in 1974: a systematic review and meta-analysis. BMC medicine, 18(1):233, 2020.

[82] Vincent Kayina, Samuel Kyobe, Fred A Katabazi, Edgar Kigozi, Moses Okee, Beatrice Odongkara, Harriet M Babikako, Christopher C Whalen, Moses L Joloba, Philippa M Musoke, et al. Pertussis prevalence and its determinants among children with persistent cough in urban uganda. PLoS One, 10(4):e0123240, 2015.

[83] Nasiha Soofie, Marta C Nunes, Prudence Kgagudi, Nadia Van Niekerk, Tselane Makgobo, Yasmeen Agosti, Cleopas Hwinya, Jayani Pathirana, and Shabir A Madhi. The burden of pertussis hospitalization in hiv-exposed and hiv-unexposed south african infants. Clinical Infectious Diseases, 63(suppl 4):S165–S173, 2016.

[84] Michiel van Boven, Neil M Ferguson, and Annelies van Rie. Unveiling the burden of pertussis. Trends in Microbiology, 12(3):116–119, 03 2004. doi: 10.1016/j.tim.2004.01.002.

[85] Sophal Ear. Towards effective emerging infectious disease surveillance: evidence from the politics of influenza in Cambodia, Indonesia, and Mexico. Politics and the Life Sciences, 33(1):69–78, 2014.

[86] R Core Team. R: A Language and Environment for Statistical Computing. R Foundation for Statistical Computing, 2022. URL https://www.R-project.org.

[87] Simon N Wood. Generalized additive models: an introduction with R. chapman and hall/CRC, 2017.

[88] Russell V. Lenth. emmeans: Estimated Marginal Means, aka Least-Squares Means, 2024. URL https://CRAN.R-project.org/package=emmeans. R package version 1.10.3.

[89] Christopher Jackson. flexsurv: A platform for parametric survival modeling in R. Journal of Statistical Software, 70 (8):1–33, 2016. doi: 10.18637/jss.v070.i08.

